# Hydroxychloroquine For Prophylaxis Of COVID-19 In Health Workers: A Randomized Clinical Trial

**DOI:** 10.1101/2021.05.14.21257059

**Authors:** Jorge Rojas-Serrano, Angelica Margarita Portillo-Vásquez Ireri Thirion-Romero, Joel Vázquez-Pérez, Fidencio Mejía-Nepomuceno Alejandra Ramírez-Venegas, Karla Midori Pérez-Kawabe, Rogelio Pérez-Padilla

## Abstract

Health care workers are at high risk of being infected with the severe acute respiratory syndrome coronavirus 2 (SARS-CoV2). Our aim is to evaluate the efficacy and safety of hydroxychloroquine (HCQ) for prophylaxis of COVID19 in health personnel exposed to patients infected by SARS-COV-2.

**Methods:** Double-blind randomized, placebo-controlled single center clinical trial. Included subjects were health care workers caring for severe COVD19 patients. Main outcome was time to symptomatic SARS-CoV2 infection.

**Results:** 127 subjects with a confirmed baseline negative RT-PCR SARS-CoV2 test were included in the trial, 62 assigned to HCQ and 65 to placebo. One subject (1.6%) in the HCQ group and 6 (9,2%) subjects in the placebo group developed COVID-19. (Log Rank test p = 0.09). No severe COVID19 cases were observed. The study was suspended because of a refusal to participate and losses to follow up after several trials reported lack of effectiveness of hydroxychloroquine in hospitalized patients with COVID-19.

**CONCLUSION:** Although the number of symptomatic infections in health personnel was lower in the HCQ group, the difference was not statistically significant. The trial is underpowered due to the failure to complete the estimated sample size.

## INTRODUCTION

Since the beginning of coronavirus disease 2019 (COVID-19) pandemic, it was evident that health care workers have a high risk of being infected with the severe acute respiratory syndrome coronavirus 2 (SARS-CoV2)^1^. In China^2^, as early as March 2020, at least 3300 health care workers had been infected, and in Italy^3^, about 20% of health care workers attending COVID-19 patients were infected. The Americas, as recognized by the director of the Pan American Health Organization (PAHO)^4^, has the highest number of infected health workers in the world and México, the highest death toll^5^. The high infection and death rate in health workers may be explained by the difficulty to supply personal protective equipment (PPE) and the inherent high exposure to SARS-CoV2 that health care workers working in the frontlines of the COVID19 pandemic have on daily basis^6^. Illness in health personnel is a serious setback in health emergencies, as it limits the possibility of care, weakens the personnel, and reduces their motivation. In addition, stress and fatigue are intense, because of difficult decisions, long working hours, and risk of infection for them and their families^7^. COVID-19 infection of health care workers could be associated with absenteeism and limited human resources to manage the pandemic^8^.

In this context, the evaluation of prophylaxis strategies against COVID19 in health care workers is an urgent need^9^. Although the first effective vaccines against COVD19 have been approved^10^, there is still a long way to go to vaccinate all health care workers. Anti-malarial drugs, have been proposed as possible therapeutic and prophylactic agents against COVID19 due to the antiviral in vitro activity of chloroquine^11^ and indeed, the in vitro activity of HCQ against SARS-CoV2 is superior to cloroquine^12^. Moreover, anti-malarial agents have immunomodulatory effects that may have a clinical benefit in COVID19 patients^13^. HCQ is a well-known drug with a good safety profile, low cost and affordable^14^. The aim of our study was to evaluate the efficacy and safety of HCQ for the prophylaxis of COVID19 in health care workers highly exposed to SARS-CoV2.

## METHODS

### Trial design

This was a double-blind, randomized, head to head placebo-controlled clinical trial, held at the National Institute of Respiratory Diseases (INER) of Mexico, a public national referral center for respiratory diseases and a main teaching center and research facility for respiratory diseases. The trial was designed and conducted by the authors. The protocol was approved by the institution’
ss review board, (C14-20) and by the COFEPRIS, the Mexican drug regulatory agency. This trail was registered in clinicaltrials.gov (number NCT04318015), This report follows the CONSORT guidelines^15^.

### Eligible subjects

Included subjects were health care workers (nurses, nursing aides, cleaning staff, orderlies, respiratory therapists and physicians) 18 years old or older, with high-risk exposure to SARS-Cov-2 as they were taking care of hospitalized patients with COVID19.. At baseline, included subjects had to be asymptomatic with a negative PCR-RT SARS-CoV2 test. Exclusion criteria were previous SARS-CoV2 infection, being allergic to hydroxychloroquine, being actual consumers of hydroxychloroquine or chloroquine (a 30 day wash out period was allowed), a weight < 50 kg, pregnancy and nursing mothers. Other exclusion criteria were tamoxifen current users, history of chronic liver disease (Child-Pugh B or C) or chronic renal disease with a glomerular filtration rate ≤ 30 ml/min. All personal in direct contact with COVID19 patients received personal protection equipment (PPE) and adequate training in the use of it. Written informed consent was obtained from all participants before study entry.

### Randomization

Eligible patients were randomized centrally, utilizing a dedicated software (http://www.randomization.com) and the results were utilized to label flasks containing 20 tablets (200 mg each) of the experimental drug and the identically appearing and packed sucrose placebo, with indications to take 1 every day for 60 days (up to three boxes containing 20 tablets of 200 mg). The flasks were given to participants each 20 days. The placebo group received an identical placebo for 60 days. Recruiters, trial team and the evaluators of follow up condition were blinded to group assignment.

### Outcomes

The primary outcome considered was the time to a symptomatic respiratory infection (cough, pharyngitis, coryza, runny nose, myalgia, arthralgia, fever or anosmia) with a positive test for SARS-CoV2 by RT-PCR over a 60-day period. Secondary endpoints were the proportion of individuals requiring hospitalization for severe COVID19, the number of days absent from work and the incidence of safety endpoints: adverse events (AE), AE leading to discontinuation and severe adverse events (SAE).

As secondary outcomes, we included the proportion of individuals with symptomatic infection by other viruses confirmed by RT-PCR in pharyngeal and/or nasopharyngeal specimen, the proportion of participants with Influenza-like illness (ILI), the proportion of individuals with missing days to work due to respiratory disease, the number of days absent from work, the proportion of participants requiring hospitalization rate for severe COVID-19.

### Procedures

At baseline evaluation, a nasopharyngeal swab sample was taken for a RT-PCR for SARS-CoV2 (Berlin Protocol, with our laboratories standardized by the Mexican National Reference Laboratory INDRE^16^). The result of the RT-PCR-was available 24 hours after the nasopharyngeal sample. Also, a complete physical examination, electrocardiogram, a general blood testing including blood cell count, blood chemistry including glucose, urea and creatinine, hepatic enzymes were done. All women in reproductive age not using a permanent contraceptive method were asked to perform a urine pregnancy test (CERTUM diagnostics, Kabla diagnosticos, Mexico).

After the baseline nasopharyngeal test, included subjects were randomized. In case that a subject had a positive SARS-CoV2 RT-PCR test, the subject was excluded from the analysis. At day 30 and 60 after treatment initiation, the same physical examination was repeated. Serum samples were taken and stored at each visit. Also, a RT-PCR for SARS-CoV2 and the pregnancy tests were repeated. All subjects were asked to report their symptoms daily via an online survey, with reminders and links being sent with a popular phone application (WhatsApp) to the phone number provided in the baseline evaluation. Subjects were also encouraged to contact the research team directly in case of a probable COVID-19 or adverse event symptom. A new RT-PCR for SARS-COV-2 was performed if subjects presented with COVID19 symptoms.

### Detection of anti-SARS-CoV-2 antibodies in serum samples

Total antibodies anti-SARS-CoV-2 were determined by ELISA using Elecsys Anti-SARS-CoV-2 immunoassay (Roche Diagnostics International Ltd, Rotkreuz, Switzerland). The immunoassay utilizes a double-antigen sandwich test principle and a recombinant protein representing the nucleocapsid antigen for the determination of antibodies (including both IgA and IgG) to SARS-CoV-2. Assay results were interpreted as follows: cutoff index, <1.0 for samples that were nonreactive/negative for anti-SARS-CoV-2 antibodies; cutoff index, ≥1.0 for samples that were reactive/positive for anti-SARS-CoV-2 antibodies.

Neutralizing antibodies were detected by ELISA using GenScript SARS-CoV-2 Surrogate Virus Neutralization Test (sVNT) Kit. Detect circulating neutralizing antibodies against SARS-CoV-2 that block the interaction between the receptor binding domain of the viral spike glycoprotein (RBD) with the ACE2 cell surface receptor. The assay detects any antibodies in serum and plasma that neutralize the RBD-ACE2 interaction, the test is both species and isotype independent. We use dilution 1:20 of each serum and inhibition of neutralization were calculated as follow: Inhibition = (1 - OD value of Sample/OD value of Negative Control) × 100%.

### Sample size

Th sample size calculation was estimated according the primary objective of the study, the time to a symptomatic SARS-CoV2 infection, assuming a 20% rate of infection in control group, as reported from Italy in February^3^, vs a 10% in the experimental group (10% reduction). Based on the time to an event analysis using a Log Rank test, we estimated a total of 400 subjects to be randomized, 200 per group. An interim analysis was planned, upon completing half of the sample. In mid-July the rhythm of recruitment was reduced drastically, with some already recruited subjects asking to leave the study, coinciding with reports both in scientific and massive media that several large trials evaluating hydroxychloroquine were suspended, due to a lack of benefit ^17–19^. We considered unlikely to complete the proposed sample size and therefore decided to suspend recruitment.

### Statistical analysis

Descriptive statistics of the studied population was according to the type of variable and distribution. All analysis were performed as intention to treat. The Kaplan-Meier method was used to estimate the time to symptomatic COVID-19 disease between the placebo and intervention groups, and the survival functions between the groups was compared using the Log-Rank test. To analyze the secondary outcomes, Pearson’s chi2 was used. In the original statistical analysis plan, a multivariate Cox regression analysis was considered to adjust for possible confounders, nevertheless, due to the lower recruitment of the trial, this analysis was not done. α was set at 5%, all analysis were two sided. Stata v 15 was used to perform all analysis.

## RESULTS

From April 21 to July 15 of 2020, 130 health care workers were randomized. Three subjects were excluded due to a positive RT-PCR SARS-CoV2 test. Finally, 127 subjects with a confirmed baseline negative RT-PCR SARS-CoV2 test were included in the trial, as shown in Figure 1. As described in Table 1, included subjects were young (overall median age 31.5 (26.7 – 40.3) and the prevalence of comorbidities excluding alcoholism and smoking was low (12.6%). Sixty-two subjects were randomized to receive hydroxychloroquine and 65 subjects received placebo. Subjects receiving hydroxychloroquine had a higher proportion of male subjects (57.4 % Vs. 35.4%, p=0.09). Also, patients randomized to receive hydroxychloroquine were using more medications, mainly over the counter NSAIDS used to relief symptoms (headache) attributed to the use of PPE by the subjects.

**Figure 1.**
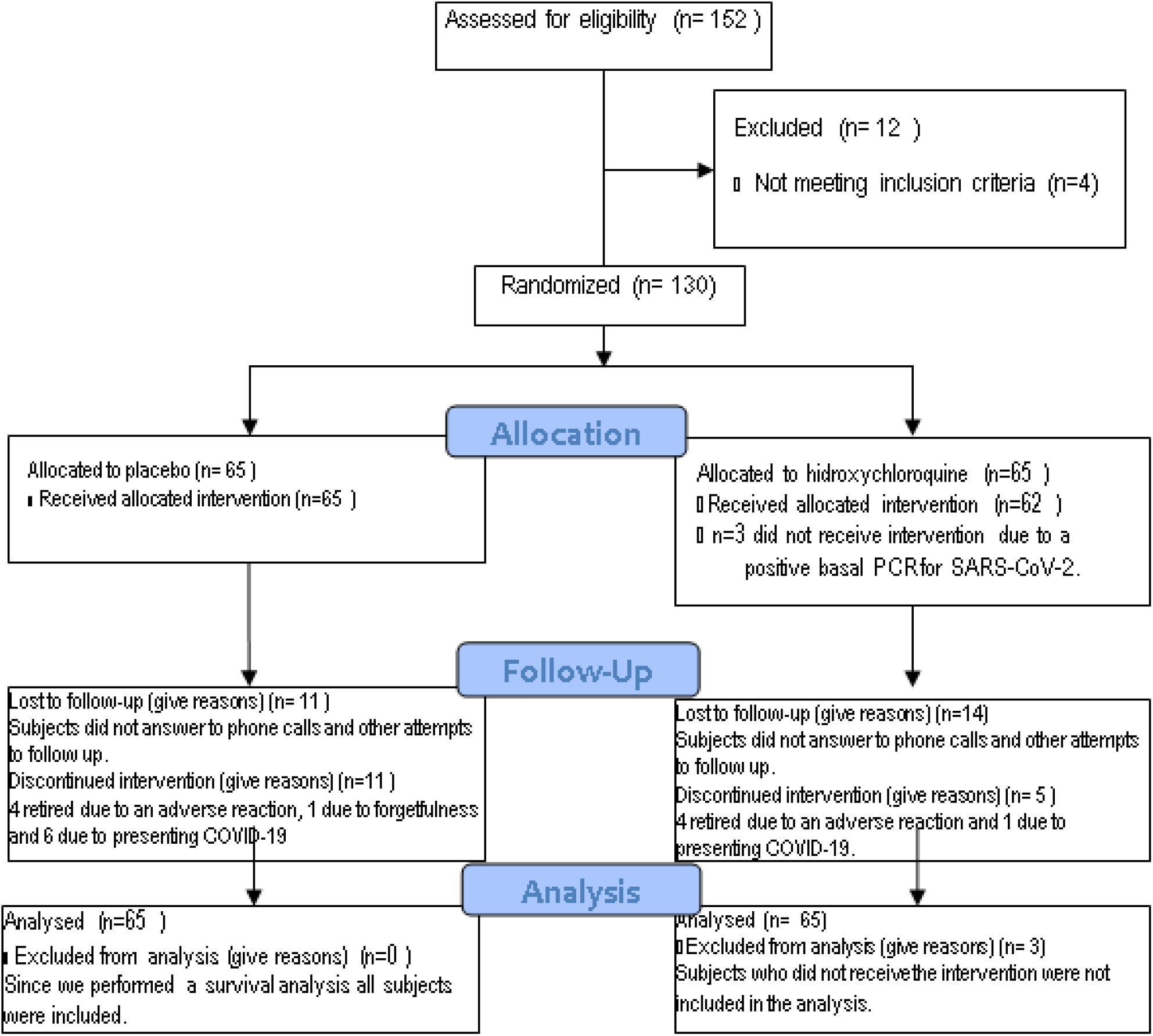
CONSORT flow diagram for the study.

**Figure 1.**
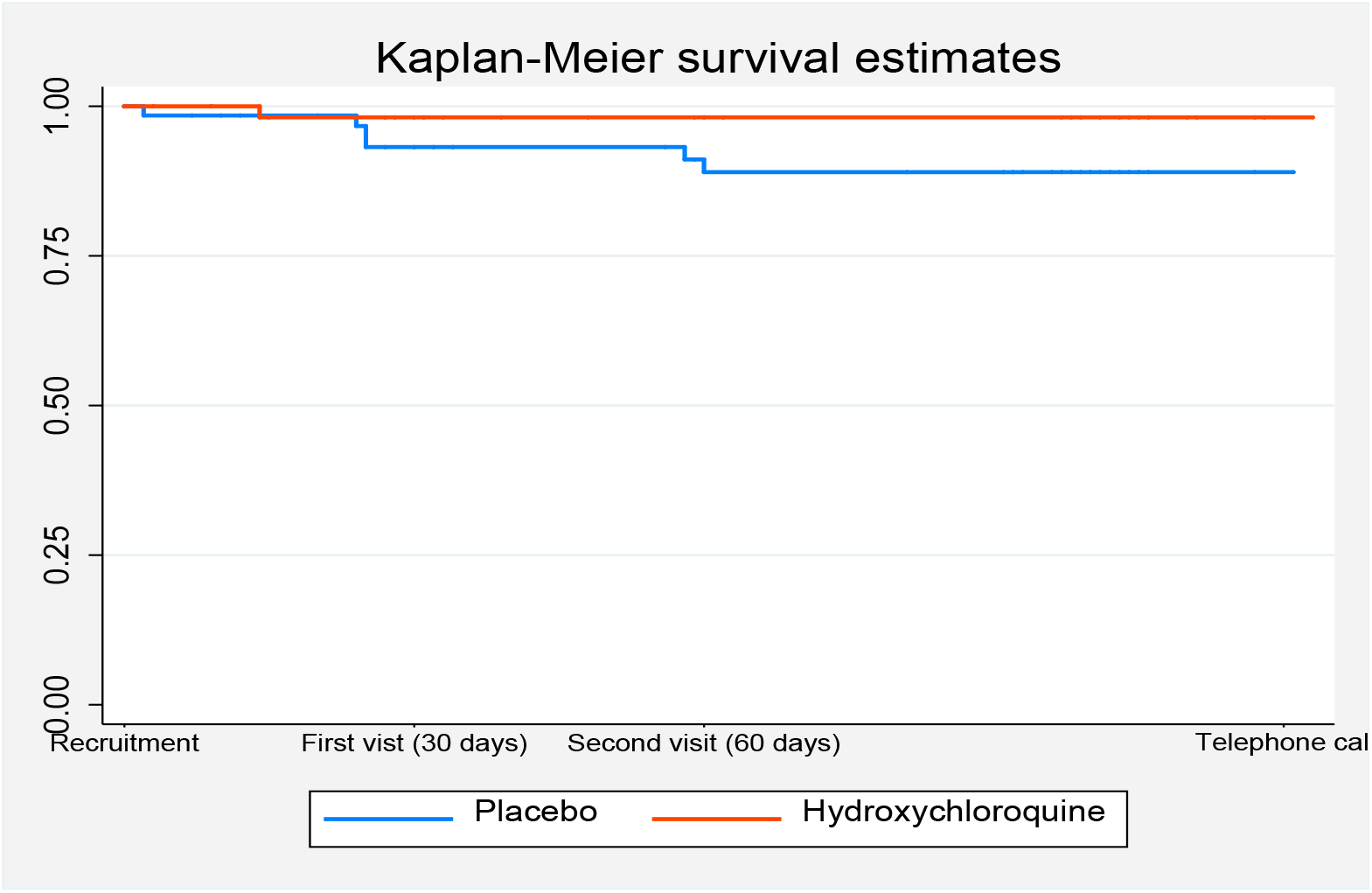
Kaplan-Meier survival curve 6 subjects in the placebo group developed symptomatic SARS-CoV2 infection, and 1 in the HCQ developed symptomatic SARS-CoV2 infection (Log Rank test p=0.09).

**TABLE 1.** CHARACTERISTICS OF THE PARTICIPANTS

|  | Placebo<br>n=65 | HCQ<br>n=62 | All subjects<br>n=127 | p |
| --- | --- | --- | --- | --- |
| Age | 31.9 (27.2 - 43.7) | 31.0 (26.4 - 39) | 31.1 (26.6 - 40.3) | 0.38 |
| Male gender (%) | 23 (35.4%) | 33 (57.4%) | 56 (42.52%) | 0.096 |
| Days of exposition to COVID-19 | 47 (28 - 64) | 40 (26 - 64) | 41 (26 - 64) | 0.52 |
| Profession (% phsycians or nurses) | 33 (50.8%) | 31 (50%) | 64 (50.39%) | 0.73 |
| Number with night shifts | 17 (26.2%) | 24 (35.5%) | 39 (30.7%) | 0.19 |
| Working at another medical center | 18 (27.7%) | 16 (25.8%) | 34 (26.8%) | 0.81 |
| Presence of concomitant disease | 7 (10.8%) | 9 (14.5%) | 16 (12.6%) | 0.52 |
| Previous or active smoking (%) | 23 (35.4%) | 20 (32.3) | 43 (33.9%) | 0.71 |
| Number reporting > 1 alcohol drink a week (%) | 8 (12.5%) | 12 (19.4%) | 20 (15.9%) | 0.29 |
| Body mass index (kg/m<sup>2</sup>) | 27.2 (4.6) | 26.7 (3.9) | 27.0 (4.3) | 0.49 |
| Obesity (%) | 15 (23.1%) | 9 (11.3%) | 24 (18.5%) | 0.079 |
| Use of other medications | 30 (46.2%) | 43 (69.4%) | 73 (57.5%) | 0.008 |

### Primary outcome results

One subject (1.6 %) in the HCQ group and 6 (9.2%) subjects in the placebo arm of the study presented COVID19 during follow-up (Log Rank test p=0.09, Figure 2.) No severe COVID-19 cases were observed, and none of the confirmed subjects needed hospitalization. All COVID-19 cases were confirmed with a positive RT-PCR SARS-CoV2 test.

Also, Cox regression models were fit to know the impact of the main secondary variables, not finding any significant risk for age, gender, days of exposition to SARS-CoV-2, time of shift, profession, or presence or a concomitant disease. These results are presented in Table 2 and Table 1 in the supplementary material.

**TABLE 2:**
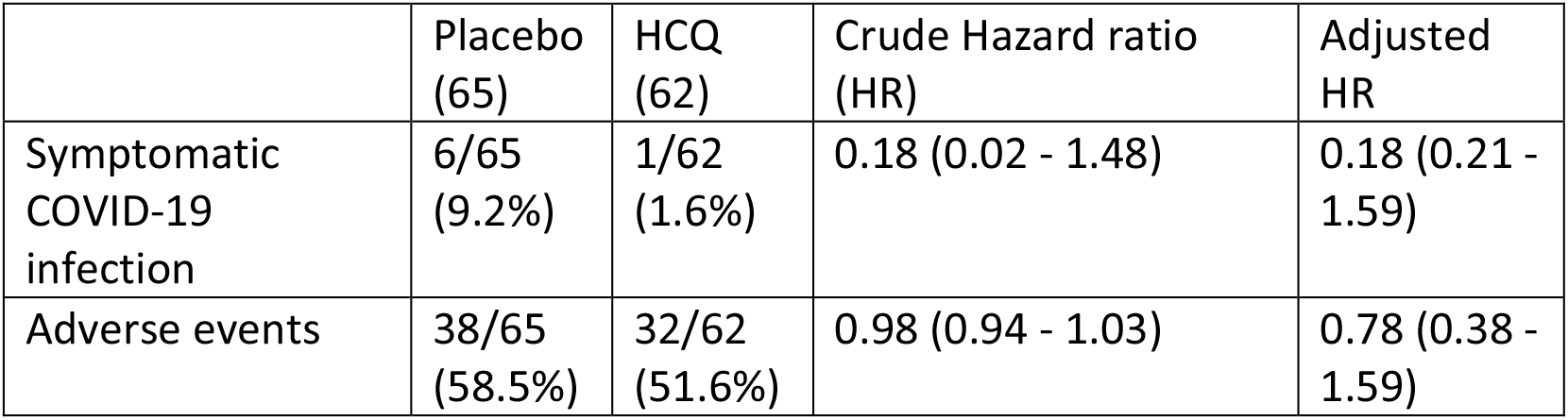
MAIN OUTCOMES

|  | Placebo<br>(65) | HCQ<br>(62) | Crude Hazard ratio<br>(HR) | Adjusted<br>HR |
| --- | --- | --- | --- | --- |
| Symptomatic<br>COVID-19<br>infection | 6/65<br>(9.2%) | 1/62<br>(1.6%) | 0.18 (0.02 - 1.48) | 0.18 (0.21 - 1.59) |
| Adverse events | 38/65<br>(58.5%) | 32/62<br>(51.6%) | 0.98 (0.94 - 1.03) | 0.78 (0.38 - 1.59) |

### Safety

One severe adverse event in the placebo group was reported (renoureteral colic). Eight participants (4 from the HCQ group) retired from the study citing an adverse reaction as the reason. In the first visit, 40 (20 placebo and 20 HCQ) out of 101 subjects reported an adverse effect, being the most common diarrhea (25 subjects), headache (22 subjects) and nausea (9 subjects). In the second visit, out of 79 subjects, 44 reported adverse effects (25 HCQ and 19 placebo, difference not significant) reported any adverse effect, with headache (21 subjects) and diarrhea (11 subjects) as the most common, all patients reporting diarrhea and headache as an adverse event had a negative PCR-RT for SARS-CoV2. There were not medically relevant abnormal results in the laboratory studies solicited throughout the study, with also not significant differences between the interventions. There were also not important differences in weight and arterial pressure. The electrocardiogram was evaluated via QT corrected for heart rate (QTc) with no important changes in the follow up visits or between groups. The details of the studies and rest of the adverse effects across the intervention groups can be seen in the supplementary material.

### Adherence to treatment

Subjects were asked at the follow-up visits to hand over the pillbox, and the remaining pills were counted and reported. 53% handed over the pillbox in the first visit and 51.9% in the second visit, with a median of 0 (0 – 2) pills remaining. In subjective adherence, most patients who attended the follow up visits reported taking the pills every day of the week, with 23.8% (first visit) and 21.3% (second visit) reporting not taking them one day, 6.9% and 6.3% not taking them two days, and 4.9% and 8.8% not taking them 3 days a week. There were not significant differences in adherence between the interventions. The most common cause for not taking the pills was forgetfulness. Thirty subjects were lost for the first follow-up visit, and 20 from the first follow up visit to the second one.

### Antibodies

Four subjects were reactive to antibodies being negative in the PCR test, 3 in the HCQ group and one in the placebo groups. Two of the subjects who received HCQ and the one assigned to placebo were reactive since the basal visit, and the rest was reactive in the first follow-up visit.

## DISCUSSION

In our study, a double-blind parallel group, randomized control trial, the time to a symptomatic SARS-CoV2 infection was delayed in the experimental group, nevertheless, this difference was not statistically significant compared with the placebo group (Log Rank test, p = 0.09, HR: 0.18 (95 % CI: 0.02 – 1.48). The results of this trial are underpowered, we were not able to fulfill the planed sample size and moreover, the observed incidence of SARS-CoV2 symptomatic infections were less than those estimated when calculating the sample size.

HCQ is a well-known drug, with a good safety profile, and has in vitro antiviral SARS CoV2 activity. HCQ is completely and rapidly absorbed after oral ingestion, nevertheless, plasma levels increase gradually and equilibrate after 3 to 4 months. HCQ achieves high concentrations in lungs. So, at the beginning of the COVD19 pandemic, HCQ was an obvious drug candidate to be evaluated in clinical trials for the treatment of severe COVID19 patients, or for the prophylaxis of COVID19 in subjects highly exposed to SARS-CoV2. Our trial is one of the several trials that have evaluated the efficacy and security of HCQ. Other trials with the same drug and population have found a lower number of infections in the HCQ groups, but without statistical significance in the analysis^20–22^. The doses and day of application vary considerably in the different essays, in general lower or for shorter time than ours, may have had an impact in the results and that some doses or days of therapy are insufficient to prevent infections. As HCQ plasma levels equilibrate after 3-4 months^23^, it is possible that a therapy for 60 days or less days, is insufficient to achieve steady-state concentrations or concentrations too low for prevention.

The number of infections in our participants was lower than expected^3^, since only seven out of 127 subjects (5.5%) were infected. This may be due to a better use of PPE and a lower incidence of disease^24^ when compared to that reported at the beginning of the pandemic in China and Italy, results that were the basis of our sample size estimation.

The population recruited were young subjects with low risk of complications, due to a Mexican policy of sending home older health care workers and those with comorbidities^25^. We are unable to exclude asymptomatic infections with SARS-CoV2 in some of our participants. Another characteristic of the population of this trial, is that more women were randomized to the HCQ group, and women seem to have a more benign course of COVID19^26^, but it is unknown if this may have affected the frequency of symptomatic COVID-19.

The eight people who retired from the study citing adverse effects were equally distributed between the randomization groups, and the reported symptoms were mostly gastrointestinal, and may point to a different origin of these, such as general anxiety (mental health was not evaluated during the study), use of PPE or changes in diet. Even though gastrointestinal adverse effects are common with HCQ^27^, the equal distribution along the randomization group and the low effect size for HCQ (0.138) don’t seem to point to an effect of HCQ. We had only one severe adverse effect, being one not known as related to HCQ.

The adherence to the intervention was good in general, but there were a high number of people lost to follow up. The reports of suspension of studies of HCQ in COVID-19 patients began with the publication of an article in Lancet (later retracted) that reported severe adverse effects and low efficacy for treatment^17^. This, and later reports of other suspensions^18,19^, undermined the general population’s trust in HCQ, since the reports were made public in general media^28^. Our subjects had several doubts about continuing the intervention, and some left the study due to their lack of trust. Information of these suspended trials traveled by newspapers and media^29^, and reached the widespread population with a great impact, even before a proper peer-reviewed publication was available and analyzed, because of the considerable prestige and importance of the institutions responsible for the trials. In addition, the participants and potential participants worked in centers dedicated to the care of COVID-19 and were well informed of developments in the treatment trials for COVID-19. Dozens of trials including HCQ as treatment were registered in Clinical Trials^20^, and it is likely that several are going to end up short, as ours did. Nonetheless, and fortunately, information can be compiled later in meta-analyses and systematic analyses. As a consequence, the trial was suspended, did not complete the planned sample size, and ended up underpowered.

We could not exclude at the baseline subjects positive to SARS-CoV2 antibodies due to lack of the antibody test at that moment, so 3 subjects previously exposed to SARS-CoV2 were included in the trial. Duration of planned prophylaxis for 2 months was based on the observed duration of the first outbreak in China^9^, but the outbreak was longer in many countries, included México and was insufficient in retrospect, but the trial could not continue recruiting.

Side effects of HCQ have been emphasized, but the population studied had in general a low risk for side effects, the dose of HCQ was as low as that used by rheumatic patients for years without a loading dose, and risks can be further reduced with proper follow-up keeping track of the QTc segment and utilizing instruments such as the multivariable Tisdale’
ss scale score to predict individuals at higher risk of QTc prolongation and its complications,

## CONCLUSION

In summary, no beneficial effect or significant harm could be demonstrated in our randomized controlled trial including 127 participants, using relatively low doses of HCQ compared with placebo in health personnel. However, the study was stopped early and likely was underpowered for finding a statistically and clinically important difference in the primary outcome.

## Data Availability

We are making our dataset available in the link below, under Creative Commons license. The protocol is the one registered at clinicaltrials.gov.

https://figshare.com/s/04ea689e248e1a52fe2c

https://clinicaltrials.gov/ct2/show/study/NCT04318015?term=phydra&draw=2&rank=1

## ACKNOWLEDGMENTS: FUNDING

Support for the trial was received from the Instituto Nacional de Enfermedades Respiratorias (INER), from CONACYT (national council of science and technology) and by SANOFI-AVENTIS through an investigator-sponsored trial. Neither the INER, CONACYT or SANOFI AVENTIS had role in the des ign of the study, collection of data, analysis, results interpretation, the writing of the final report and the decision to submit the article to publication.

