## Supplemental tables 1 to 3 for "Hydroxychloroquine For Prophylaxis Of COVID-19 In Health Workers: A Randomized Clinical Trial"

**On behalf of the** RESEARCH GROUP ON HYDROXYCHLOROQUINE FOR COVID-19 also formed by Dr. Cristobal Guadarrama, Dr. Carmen Margarita Hernández Cárdenas, Dr. Luis Felipe Jurado Camacho Dr. Sebastián Rodríguez Llamazares, Dra. Alejandra Ramírez Venegas

ACKNOWLEDGMENTS: FUNDING
Support for the trial was received from the Instituto Nacional de Enfermedades Respiratorias (INER), from CONACYT (national council of science and technology) and by SANOFI-AVENTIS through an investigator-sponsored trial. Neither the INER, CONACYT or SANOFI AVENTIS had role in the des ign of the study, collection of data, analysis, results interpretation, the writing of the final report and the decision to submit the article to publication.

**SUMMARY**

Health care workers are at high risk of being infected with the severe acute respiratory syndrome coronavirus 2 (SARS-CoV2). Our aim is to evaluate the efficacy and safety of hydroxychloroquine (HCQ) for prophylaxis of COVID19 in health personnel exposed to patients infected by SARS-COV-2.

**Methods:** Double-blind randomized, placebo-controlled single center clinical trial. Included subjects were health care workers caring for severe COVD19 patients. Main outcome was time to symptomatic SARS-CoV2 infection.

**Results:** 127 subjects with a confirmed baseline negative RT-PCR SARS-CoV2 test were included in the trial, 62 assigned to HCQ and 65 to placebo. One subject (1.6%) in the HCQ group and 6 (9,2%) subjects in the placebo group developed COVID-19. (Log Rank test p = 0.09). No severe COVID19 cases were observed. The study was suspended because of a refusal to participate and losses to follow up after several trials reported lack of effectiveness of hydroxychloroquine in hospitalized patients with COVID-19.

**CONCLUSION**

Although the number of symptomatic infections in health personnel was lower in the HCQ group, the difference was not statistically significant. The trial is underpowered due to the failure to complete the estimated sample size.

**Key words:** COVID-19, health workers, prophylaxis, hydroxychloroquine,

**SUPLEMENTARY MATERIAL**.

TABLE S1: MULTIVARIATE MODELS

| | Simple regression, unadjusted models | | | Multiple regression, adjusted models | | |
|---|---|---|---|---|---|---|
| | Hazard ratio | 95% confidence interval | p | Hazard ratio | 95% confidence interval | p |
| Assignated intervention | 0.18 | 0.021 - 1.48 | 0.11 | 0.18 | 0.21 - 1.59 | 0.12 |
| Age at recruitment | 0.94 | 0.85 - 1.05 | 0.27 | 0.94 | 0.84 - 1.05 | 0.26 |
| Gender | 0.99 | 0.222 - 4.43 | 0.99 | 1.39 | 0.26 - 7.45 | 0.70 |
| Presence of concomitant disease | 1.14 | 0.255 - 5.094 | 0.86 | 0.66 | 0.11 - 4.0 | 0.65 |
| Body mass index | 1.23 | 1.06 - 1.42 | 0.006 | 1.22 | 1.05 - 1.42 | 0.01 |
| Profession (Nurse or physician/other) | 2.14 | 0.42 - 11.0 | 0.36 | 1.99 | 0.37 - 10.67 | 0.42 |
| Shift schedule (not night/night) | 1.64 | 0.37 - 7.35 | 0.52 | 1.57 | 0.29 - 8.24 | 0.59 |
| Days of exposition to COVID-19 | 0.995 | 0.97 - 1.03 | 0.73 | 1 | 0.96 - 1.04 | 0.97 |

TABLE S2.

| | Visit 1 (All) (n=30) | Visit 2 (All) (n=100) | Visit 3 (All n=80) | Visit 1 (P) (n=65) | Visit 1 (H) (n=65) | Visit 2 (P) (n=49) | Visit 2 (H) (n=51) | Visit 3 (P) (n=40) | Visit 3 (H) (n=40) | |
|---|---|---|---|---|---|---|---|---|---|---|
| Leukocytes | 6.4 (5.5 - 7.9) | 6.4 (5.6 - 7.3) | 6.5 (5.75 - 7.41) | 6.4 (5.4 - 8.1) | 6.5 (5.5 - 7.4) | 6.4 (5.6 - 7.4) | 6.4 (5.6 - 7.1) | 6.6 (5.75 - 7.6) | 6.35 (5.75 - 7.25) | 0.38 |
| Plateletes | 248 (220 - 283) | 250 (225 - 278) | 257.5 (233 - 282.5) | 248 (226 - 274) | 248 (202 - 283) | 254 (232 - 284) | 244 (221 - 275) | 253.5 (236 - 283.5) | 259 (221.5 - 280) | 0.24 |
| Lymphocytes | 2 (1.6 - 2.4) | 2 (1.65 - 2.5) | 1.95 (1.6 - 2.5) | 2 (1.6 - 2.5) | 2 (1.6 - 2.3) | 2 (1.7 - 2.5) | 2 (1.5 - 2.5) | 1.95 (1.5 - 2.35) | 1.95 (1.65 - 2.5) | 0.45 |
| Neutrophils | 3.6 (3.1 - 4.6) | 3.7 (3 - 4.65) | 3.7 (3.1 - 4.8) | 3.8 (3.1 - 4.7) | 3.6 (2.9 - 4.5) | 3.7 (3 - 4.7) | 3.7 (3.1 - 4.5) | 3.9 (3.2 - 4.8) | 3.45 (3.05 - 4.75) | 0.27 |
| Eosinophils | 0.1 (0.1 - 0.2) | 0.1 (0.1 - 0.2) | 0.1 (0.1 - 0.2) | 0.1 (0.1 - 0.2) | 0.1 (0.1 - 0.2) | 0.1 (0.1 - 0.2) | 0.1 (0.1 - 0.2) | 0.1 (0.1 - 0.2) | 0.1 (0.1 - 0.2) | 0.51 |
| Creatinin | 0.79 (0.67 - 0.94) | 0.8 (0.692 - 0.915) | 0.775 (0.67 - 0.915) | 0.76 (0.65 - 0.88) | 0.87 (0.68 - 0.95) | 0.74 (0.66 - 0.89) | 0.84 (0.71 - 0.92) | 0.715 (0.655 - 0.815) | 0.825 (0.72 - 0.92 | 0.83 |
| BUN | 13 (11 - 15) | 12 (11 - 15) | 13 (10 - 15) | 13 (11 - 15) | 13 (11 - 15.17) | 12 (11 - 15) | 13 (11 - 16) | 13.5 810 - 15) | 13 (11 - 15.5) | 0.91 |
| ALT | 19.5 (14 - 31) | 18.5 (14 - 24) | 18 (14 - 28) | 18 (13 - 26) | 21 (15 - 35.7) | 18 (15 - 25) | 20 (13 - 32) | 18 (14 - 27.5) | 18.5 (14 - 30) | 0.3 |
| AST | 19 (16 - 24) | 19 (17 - 24) | 19 (16 - 23) | 18 (15 - 22.3) | 20 (18 - 27) | 18 (16 - 22) | 19 (17 - 25) | 19 (16 - 22.5) | 20 (16 - 23.5) | 0.299 |
| Total bilirrubin | 0.53 (0.41 - 0.79 | 0.54 (0.41 - 0.73) | 0.52 (0.385 - 0.66) | 0.53 (0.42 - 0.71) | 0.53 (0.41 - 0.66) | 00.52 (0.42 - 0.73) | 0.57 (0.41 - 0.72) | 0.455 (0.37 - 0.635) | 0.525 0.395 - 0.74) | 0.46 |
| Direct bilirrubin | 0.1 (0.08 - 0.13) | 0.1 (0.08 - 0.135) | 0.1 (0.075 - 0.14) | 0.1 (0.08 - 0.13) | 0.1 (0.08 - 0.13) | 0.1 (0.07 - 0.13) | 0.11 (0.08 - 0.14) | 0.09 (0.07 - 0.12) | 0.12 (0.085 - 0.15) | 0.28 |
| Systolic blood pressure | 112 (104 - 121) | 111 (103 - 120) | 110.5 (103.5 - 120) | 112 (104 - 122) | 112 (106 - 119) | 111 (102 - 123) | 112 (103 - 118) | 110 (103 - 117.5) | 113.5 (103.5 - 120) | 0.63 |
| Diastolic blood pressure | 71 (67 - 77) | 70 (64 - 75) | 71 (66.5 - 75) | 72 (67 - 79) | 71 (66 - 76) | 69 (63 - 75) | 72 (64 - 76) | 72 (68 - 77.5) | 70 (65 - 74.5) | 0.049 |
| Heart rate | 74 (67 - 81) | 77 (70 - 85) | 77.5 (69 - 84) | 75 (67 - 81) | 74 (66 - 78) | 79 (70 - 86) | 76 (69 - 82) | 80 (71 86.5) | 75 (66.5 - 81.5) | 0.97 |
| Temperature | 36.5 (36.3 - 36.8) | 36.5 (36.3 - 36.7) | 36.5 (36.4 - 36.7) | 36.6 (36.3 -36.7) | 36.5 (36.3 - 36.8) | 36.5 (36.4 - 36.7) | 36.4 (36.3 - 36.7) | 36.5 (36.4 - 36.7) | 36.5 (36.4 - 36.7) | 0.67 |
| RR | 22 (19 - 23) | 22 (19 - 23) | 20 (19 - 22) | 22 (19 - 23) | 22 (19 - 22) | 22 (19 - 23) | 22 (19 - 23) | 21 (19 - 22) | 20 (19 - 22) | 0.43 |
| QTc | 407 (390 - 424) | 409 (389 - 429) | 412 (390 - 427) | 414.5 (389 - 426.5) | 403 (390 - 420 | 412 (386 - 433) | 404 (389 - 428) | 413.5 (381 - 427) | 410 (394.5 424.5) | 0.34 |
| Weight | 70.1 (60 - 80.7) | 69 (60.2 - 78) | 68.9 (60.7 - 77.65) | 66.9 (59.9 - 80.8) | 72.7 (60 - 80) | 67.05 (60.2 - 76.8) | 70 (60 - 80.3) | 67.8 (61.2 - 76) | 70.8 (60.45 - 79.3) | 0.89 |
| Height | 161 (155 - 168) | 161 (155 - 168) | 160.5 (154 - 168.5) | 159 (152 - 166) | 163 (157 - 168) | 159 (152 - 164) | 163 (157 - 170) | 157 (135 - 164) | 163.5 (157 - 171) | 0.052 |
| BMI | 26.8 (24.4 - 28.89) | 26.4 (23.9 - 28.2) | 26.1 (23.85 - 28.2) | 26.85 (24.2 - 29.65) | 26.2 (24.6 - 28.5) | 26.65 (24.57 - 28.8) | 26.1 (237 - 28.1) | 26.5 (23.78 - 28.95) | 25.95 (23.85 27.7 | 0.14 |

Supplementary table 2. Findings in laboratory studies and clinical examination throughout the study.

TABLE S3

| | First follow up visit | | | Second follow up visit | | |
|---|---|---|---|---|---|---|
| | Placebo | HCQ | p | Placebo | HCQ | p |
| Adverse effect at visit | 20 (40%) | 20 (39.22%) | 0.896 | 25 (62.5%) | 19 (47.5%) | 0.157 |
| Visual adverse effect | 4 (8%) | 1 (2%) | 0.177 | 1 (2.5%) | 0 | 0.58 |
| Lack of focus | 1 | 0 | | 0 | 0 | |
| Red spots | 1 | 0 | | 0 | 0 | |
| Blurry vision | 2 | 1 | | 1 | 0 | |
| Adverse cardiologic effects | 3 (6%) | 3 (5.88%) | 0.664 | 1 (2.5%) | 1 (2.5%) | 0.672 |
| Hypotension | 1 | 2 | | 0 | 0 | |
| Thoracic opression | 1 | 0 | | 0 | 0 | |
| Taquicardia | 1 | 1 | | 0 | 0 | |
| Agitation | 0 | 0 | | 1 | 0 | |
| Palpitations | 0 | 0 | | 0 | 1 | |
| Adverse neurological effects | 17 (34%) | 11 (21.57%) | 0.104 | 14 (35%) | 9 (22.5%) | 0.449 |
| Agitation | 0 | 0 | | 1 | 0 | |
| Dizziness | 2 | 2 | | 2 | 3 | |
| Headache | 14 | 8 | | 13 | 8 | |
| Paresthesia | 4 | 1 | | 1 | 1 | |
| Lipothymia | 0 | 1 | | 0 | 0 | |
| Adverse gastrointestinal effects | 17 (34%) | 24 (47.05%) | 0.059 | 9 (22.5%) | 8 (20%) | 0.419 |
| Vomit | 0 | 1 | | 1 | 0 | |
| Nausea | 5 | 4 | | 2 | 3 | |
| Diarrhea | 9 | 16 | | 7 | 4 | |
| Epigastralgia | 1 | 0 | | 0 | 0 | |
| Abdominal pain | 1 | 2 | | 0 | 1 | |
| Gastritis | 1 | 1 | | 0 | 1 | |
| Pyrosis | 1 | 0 | | 0 | 0 | |
| Constipation | 1 | 0 | | 0 | 0 | |
| Increased peristalsis | 0 | 1 | | 0 | 0 | |
| Abdominal distension | 0 | 2 | | 1 | 0 | |
| Hematochezia | 0 | 1 | | 0 | 0 | |
| Colitis | 0 | 0 | | 1 | 0 | |
| Adverse dermatologic effect | 8 (16%) | 3 (5.88%) | 0.092 | 8 (20%) | 1 (2.5%) | 0.037 |

| | | | | | | |
|---|---|---|---|---|---|---|
| Pruritus | 5 | 1 | | 5 | 0 | |
| Urticaria | 2 | 1 | | 0 | 1 | |
| Hyperpigmentation | 1 | 0 | | 1 | 0 | |
| Rash | 0 | 1 | | 0 | 0 | |
| Alopecia | 0 | 0 | | 1 | 0 | |
| Pustulas | 0 | 0 | | 1 | 0 | |
| Adverse audiological effect | 3 (6%) | 2 (3.92%) | 0.5 | 1 (2.5%) | 2 (5%) | 0.382 |
| Tinnnitus | 1 | 1 | | 0 | 1 | |
| Hearing loss | 1 | 0 | | 0 | 0 | |
| Vertigo | 2 | 0 | | 1 | 0 | |
| Right otalgia | 0 | 1 | | 0 | 1 | |
| Other adverse effects | 2 (4%) | 3 (5.88%) | 0.5 | 4 (10%) | 3 (7.5%) | 0.641 |
| Artralgia | 2 | 0 | | 1 | 0 | |
| Night sweating | 0 | 1 | | 0 | 1 | |
| Hyporexia | 0 | 1 | | 0 | 0 | |
| Tiredness | 0 | 1 | | 0 | 1 | |
| Odinofagia | 0 | 0 | | 1 | 0 | |
| Rhinorrea | 0 | 0 | | 1 | 0 | |
| Abnormal transvaginal bleeding | 0 | 0 | | 0 | 1 | |

Supplementary Table 3. Adverse events by intervention, first and second follow up visit.
